# Association between Positive Airway Pressure Adherence and Healthcare Costs

**DOI:** 10.1101/2021.09.10.21263396

**Authors:** Jaejin An, Henry A. Glick, Amy M. Sawyer, Jessica Arguelles, Charles J. Bae, Aiyu Chen, Brendan T. Keenan, Samuel T. Kuna, Greg Maislin, Diego R. Mazzotti, Allan I. Pack, Jiaxiao M. Shi, Alexa J. Watach, Dennis Hwang

## Abstract

**Background:** The impact of positive airway pressure (PAP) therapy for obstructive sleep apnea (OSA) on healthcare costs is uncertain.

**Research Question:** Are three-year healthcare costs associated with PAP adherence in participants from the Tele-OSA clinical trial?

**Study Design and Methods:** Participants with OSA and prescribed PAP in the Tele-OSA study were stratified into three PAP adherence groups based on usage patterns over three years: (a) high (consistently ≥ 4 hours/night); (b) moderate (2-3.9 hours/night or inconsistently ≥ 4 hours/night); (c) low (<2 hours/night). Using data from three months of the Tele-OSA trial and 33 months of post-trial follow-up, average healthcare costs (2020 US dollars) in six-month intervals were derived from electronic health records and analyzed using multivariable generalized linear models.

**Results:** Of 543 participants, 25% were categorized as having high adherence, 22% moderate adherence, and 52% low adherence to PAP therapy. Average (standard deviation) PAP use was 6.5 (1.0) hours, 3.7 (1.2) hours, and 0.5 (0.5) hours for the high, moderate, and low adherence groups, respectively. The high adherence group had the lowest average [standard error] covariate-adjusted six-month healthcare costs ($3,162 [$240]) compared with the moderate ($3,658 [$369]) and low ($4,016 [$315]) adherence groups. Significant cost savings were observed between the high and low adherence groups ($854 [95% CI $158, $1,551]); savings between moderate and low adherence were non-significant ($359 [95% CI -$459, $1,176]).

**Interpretation:** In participants with OSA, better PAP adherence was associated with significantly lower healthcare costs over three years. Findings support the importance of strategies to enhance long-term PAP adherence.

---

Obstructive sleep apnea (OSA) carries substantial public health implications given its high prevalence and associations with co-morbid medical conditions. OSA affects approximately 26% of adults between the ages of 30 and 70 years in the United States and 1 billion persons worldwide [1, 2]. Furthermore, it is strongly linked to a broad range of cardiometabolic conditions including hypertension, diabetes, arrhythmias, heart failure, stroke, acute cardiovascular events, and death [3-12]. The impact of OSA on mental health and various components of daytime functioning such as excessive daytime somnolence is also well-established resulting in increased risk of psychiatric disorders, motor vehicle accidents, and work absenteeism [13-22]. As a result, OSA is understood to directly and indirectly impact patient, societal, and healthcare costs [23-43].

The increase in health-related costs is reflected in a broad spectrum of services affecting inpatient, outpatient, emergency, and pharmacy-related costs [34, 43-45]. Conceptually, positive airway pressure (PAP) therapy should mitigate the impact of OSA on healthcare utilization; however, the overall body of evidence is limited and its results are mixed [34, 44-47]. Furthermore, recent technological advances in PAP devices have resulted in the ability to remotely monitor objective measures of PAP adherence over time. These measures are important when investigating the relationship between PAP adherence and cost because as few as 30-40% of patients have been reported to maintain substantial use after 1 year [48].

In this study, we used data from the Tele-OSA randomized clinical trial that evaluated the impact of an automated telemonitoring mechanism on improving PAP adherence [48]. The goal of this *post hoc* analysis is to evaluate the relationship between three levels of long-term PAP adherence and three-year healthcare cost.

## Methods

### Clinical Trial

Data for our analysis were drawn from the Tele-OSA study, a four-arm factorial design randomized clinical trial that enrolled participants from 2015 to 2016. The study investigated the impact of two telehealth mechanisms on PAP adherence: (a) remote delivery of an automated digital OSA educational program (Emmi Solutions, Inc), and (b) automated PAP telemonitoring with participant feedback messaging (see **e-Figure 1**). A total of 1,455 participants were enrolled in the study, of which 556 agreed to participate in the PAP trial and accept PAP therapy. Study results revealed significantly improved three-month PAP adherence (primary endpoint of the Tele-OSA study) in participants receiving telemonitoring [48]. Epidemiologic follow-up based on electronic health records was continued for an additional 33 months, for a total follow-up of three years. *Post-hoc* analysis demonstrated improved PAP use at one year with continued feedback messaging [49].

### Ethics

The Tele-OSA study and all subsequent analyses based on the study cohort were approved by the Kaiser Permanente Southern California Institutional Review Board. Participant written informed consent was waived for the entire study, including the epidemiologic follow-up, due to the minimal risk of the study interventions and data collection procedures.

### Study Population

Of the 1,455 trial enrollees, we included participants with OSA (apnea hypopnea index ≥ 5 events/hour) who were prescribed PAP. To ensure sufficient cost data for our analysis, we excluded participants without six months or longer continuous health plan eligibility during the three-year follow-up period.

### PAP Adherence

Hours of PAP use were derived from an auto-titrating device (AirSense 10; ResMed Corp) capable of wirelessly transmitting PAP data daily via a cellular signal into a cloud database (U-Sleep; ResMed Corp). We created PAP adherence groups by consolidating eligible participants from all study arms and used these data to stratify them into high, moderate, and low use groups. We considered two dimensions of PAP adherence to construct these categories—hours/night of daily use and consistency of use throughout the three years of follow-up.

The high adherence group consisted of participants who used PAP an average of ≥ 4 hours/night consistently for every six-month interval over three years. The moderate adherence group consisted of participants who either used PAP for an overall average of 2-3.9 hours/night over three years or who used PAP for ≥ 4 hours/night overall, but had at least one six-month interval with <4 hours/night of average use (i.e., inconsistent use). The low adherence group consisted of participants who used PAP an average of <2 hours/night over three years, independent of whether they had any six-month intervals with >4 hours/night.

### Healthcare Costs

The trial was performed in an integrated healthcare system that allowed access to near-comprehensive healthcare utilization information maintained in electronic health records. Healthcare costs (2020 US dollars) were derived by assigning nationally representative cost estimates to this utilization. Recorded healthcare services included sleep-related and non-sleep-related office visits, sleep-related and non-sleep-related durable medical equipment (DME), hospitalizations, other hospital visits (e.g., observation bed, same-day discharge after minimally invasive surgery), pharmacy, laboratory tests, emergency department visits, radiology visits, and phone encounters.

Current Procedural Terminology (CPT) and Healthcare Common Procedure Coding System (HCPCS) Codes were used to assign cost estimates from 2020 Medicare fee schedules for each service other than hospitalization and pharmacy [50-52]. Sleep-related CPT codes and Medicare fee schedules are shown in **eTable 1**. For pharmacy costs, we used Medicaid National Average Drug Acquisition Cost (NADAC) data to assign costs to each National Drug Code or generic name per prescription [53]. For hospitalizations, we used data from the National Inpatient Sample to derive an average cost per day for each Diagnosis Related Group code and multiplied it by participants’ lengths of stay [54]. The Consumer Price Index Medical Care component was applied to convert all hospitalization costs to 2020 US dollars [55].

### Statistical Analysis

We compared participant sociodemographic and clinical characteristics among the PAP adherence groups using analysis of variance (ANOVA) for continuous variables and Chi-square tests for categorical variables. They were also used for the evaluation of average (SD) hours/night of PAP use and healthcare costs within six-month intervals during the three years of follow-up. Trends over time were tested using non-parametric Jonckheere-Terpstra tests.

Adjusted six-month average healthcare costs by adherence group were estimated using repeated measures generalized linear models (GLM) that accounted for within-subject correlations. Covariates included age, sex, race/ethnicity, body mass index (BMI) category, Medicaid enrollment, smoking, Apnea Hypopnea Index (AHI) category (5-14.9, 15-29.9, 30 or higher), hypertension, Charlson comorbidity score, mild liver disease, pre-operative referral to sleep clinic prior to randomization, log transformed prior six-month cost, and time indicators representing the six-month interval (i.e. months 1-6, months 7-12, etc.). To address bias due to censoring (e.g., lost membership during follow-up), we applied inverse probability of censoring weights estimated by use of logistic regression [56, 57]. For the GLM model predicting total cost as well as for the models predicting subtypes of costs (e.g., hospitalization and pharmacy), we used the modified Park, Pregibon link, Pearson correlation, and modified Hosmer and Lemeshow tests to select families and links [58]. For example, the resulting recommended link for the estimation of total costs was power = -0.3 and family was gamma.

An *a priori* stratified analysis was conducted by OSA severity (AHI 5-14.9 and 15 or higher). Effect modification by OSA severity was formally tested using statistical interaction tests. Sensitivity analysis was conducted after excluding participants who were referred to sleep clinic prior to any planned surgery.

All statistical analyses were performed using SAS 9.4 (SAS Institute, Cary, NC) or Stata 15.1 (StataCorp LLC, College Station, TX). Given our single primary outcome of total costs, a *p*<0.05 was considered statistically significant.

## Results

Of the 1,455 participants that were enrolled in the Tele-OSA study, we excluded 372 participants with an AHI <5, 531 who had an AHI >5 and were not prescribed PAP therapy, and an additional 9 participants for not having at least six months of continuous eligibility. Of the remaining 543 participants, 138 (25%) were categorized as having high adherence to PAP therapy, 120 (22%) moderate adherence, and 285 (52%) low adherence.

Participant sociodemographic and clinical characteristics overall and by PAP adherence groups are shown in **Table 1**. The sample was middle aged (mean [SD] age 50.0 [12.1] years) and obese (BMI of 35.3 [7.5] kg/m^2^) on average, and 59% were male. A greater proportion of the high adherence group was comprised of whites compared with the low and moderate adherence groups. Mean AHI scores were higher for the high adherence group than for the low or moderate adherence groups, although all groups had moderate/severe disease on average. Other characteristics were similar across PAP adherence groups.

**Table 1.** Participant Sociodemographic and Clinical Characteristics by PAP Adherence.

| Characteristics | Total<br>(N=543) | Low<br>Adherence<br>(N=285) | Moderate<br>Adherence<br>(N=120) | High<br>Adherence<br>(N=138) | p-value |
| --- | --- | --- | --- | --- | --- |
| Age, mean (SD) | 50.0 (12.1) | 49.3 (12.5) | 50.9 (11.9) | 50.8 (11.6) | 0.34 |
| Male, N (%) | 321 (59) | 155 (54) | 73 (61) | 93 (67) | 0.04 |
| Race/ethnicity, N (%) |  |  |  |  |  |
| Asian Pacific Islander | 42 (8) | 23 (8) | 11 (9) | 8 (6) | 0.01 |
| Non-Hispanic Black | 47 (9) | 29 (10) | 12 (10) | 6 (4) |  |
| Hispanic | 215 (40) | 123 (43) | 48 (40) | 44 (32) |  |
| Non-Hispanic White | 228 (42) | 102 (36) | 47 (39) | 79 (57) |  |
| Other | 11 (2) | 8 (3) | 2 (2) | 1 (1) |  |
| Body Mass Index, mean (SD) | 35.3 (7.5) | 35.9 (7.4) | 35.1 (8.6) | 34.4 (6.4) | 0.15 |
| Body Mass Index Category, N (%) |  |  |  |  |  |
| Normal | 24 (4) | 11 (4) | 7 (6) | 6 (4) | 0.47 |
| Overweight | 110 (20) | 53 (19) | 29 (24) | 28 (20) |  |
| Moderately obese | 279 (51) | 147 (52) | 58 (48) | 74 (54) |  |
| Severe obese | 124 (23) | 73 (26) | 24 (20) | 27 (20) |  |
| Missing | 6 (1) | 1 (0.4) | 2 (2) | 3 (2) |  |
| Medicaid indicator, N (%) | 42 (8) | 22 (8) | 10 (8) | 10 (7) | 0.95 |
| Smoking, N (%) |  |  |  |  |  |
| Never/passive | 340 (63) | 187 (66) | 65 (54) | 88 (64) | 0.31 |
| Quit | 156 (29) | 73 (26) | 46 (38) | 37 (27) |  |
| Active | 36 (7) | 19 (7) | 7 (6) | 10 (7) |  |
| Missing | 11 (2) | 6 (2) | 2 (2) | 3 (2) |  |
| Neighborhood median household income, mean (SD) | \$77,330<br>(\$27,730) | \$76,022<br>(\$27,787) | \$79,035<br>(\$29,275) | \$78,578<br>(\$26,250) | 0.51 |
| Neighborhood % of high school or less, mean (SD) | 42.3 (15.8) | 43.6 (16.0) | 42.3 (15.4) | 39.6 (15.2) | 0.06 |
| Marital status, N (%) |  |  |  |  |  |
| Married/partner | 357 (66) | 174 (61) | 84 (70) | 99 (72) | 0.09 |
| Single/divorced/widow | 146 (27) | 90 (32) | 25 (21) | 31 (22) |  |
| Unknown | 40 (7) | 21 (7) | 11 (9) | 8 (6) |  |
| Epworth Sleepiness Scale (ESS), mean (SD) | 9.5 (5.2) | 9.5 (5.3) | 9.8 (5.2) | 9.3 (5.0) | 0.74 |
| ESS category, N (%) |  |  |  |  |  |
| <11 | 324 (60) | 167 (59) | 69 (58) | 88 (64) | 0.77 |
| 11-14.9 | 105 (19) | 55 (19) | 26 (22) | 24 (17) |  |
| 15 or higher | 103 (19) | 55 (19) | 24 (20) | 24 (17) |  |
| Missing | 11 (2) | 8 (3) | 1 (1) | 2 (1) |  |
| Functional Outcomes of Sleep Questionnaire-10), mean (SD) | 13.8 (4.5) | 13.5 (4.5) | 14.0 (4.5) | 14.2 (4.5) | 0.25 |
| Missing, N (%) | 26 (5) | 18 (6) | 11 (9) | 9 (7) |  |
| Apnea Hypopnea Index (AHI), mean (SD) | **32.0 (25.7)** | **27.7 (24.9)** | **36.6 (27.2)** | **36.7 (24.7)** | **<0.001** |
| AHI category, N (%) |  |  |  |  |  |
| 5-14.9 | **169 (31)** | **108 (38)** | **28 (23)** | **33 (24)** | **<0.001** |
| 15-29.9 | **155 (29)** | **88 (31)** | **34 (28)** | **33 (24)** |  |
| 30 or higher | **219 (40)** | **89 (31)** | **58 (48)** | **72 (52)** |  |
| Hypertension diagnosis, N (%) | 214 (39) | 109 (38) | 50 (42) | 55 (40) | 0.81 |
| Charlson Comorbidity Score (weighted), mean (SD) | 0.9 (1.3) | 0.9 (1.4) | 0.7 (1.2) | 0.9 (1.5) | 0.38 |
| Charlson Comorbidity Category, N (%) |  |  |  |  |  |
| Diabetes without complications | 112 (21) | 65 (23) | 25 (21) | 22 (16) | 0.26 |
| Diabetes with complications | 43 (8) | 25 (9) | 8 (7) | 10 (7) | 0.73 |
| Chronic pulmonary disease | 115 (21) | 66 (23) | 20 (17) | 29 (21) | 0.34 |
| Peripheral vascular disease | 32 (6) | 16 (6) | 6 (5) | 10 (7) | 0.72 |
| Renal disease | 28 (5) | 13 (5) | 3 (3) | 12 (9) | 0.06 |
| Mild liver disease | **32 (6)** | **23 (8)** | **7 (6)** | **2 (1)** | **0.03** |
| Cancer | 20 (4) | 10 (4) | 2 (2) | 8 (6) | 0.21 |
| Congestive heart failure | 8 (1) | 4 (1) | 2 (2) | 2 (1) | 0.98 |
| Cerebrovascular disease | 9 (2) | 4 (1) | 3 (3) | 2 (1) | 0.72 |
| Myocardial Infarction | 8 (1) | 1 (0.4) | 4 (3) | 3 (2) | 0.06 |
| Rheumatic disease | 4 (1) | 3 (1) | 0 (0) | 1 (1) | 0.53 |
| Paraplegia/hemiplegia | 3 (1) | 1 (0.4) | 2 (2) | 0 (0) | 0.16 |
| Metastatic Carcinoma | 3 (1) | 3 (1) | 0 (0) | 0 (0) | 0.26 |
| Moderate/severe liver disease | 1 (0.2) | 1 (0.4) | 0 (0) | 0 (0) | 0.64 |
| Sleep clinic referral due to pre-operative assessment, N (%) | **36 (7)** | **27 (9)** | **6 (5)** | **3 (2)** | **0.01** |
| Medication adherence |  |  |  |  |  |
| Antihypertensive medications, N (%) | 255 (47) | 132 (46) | 58 (48) | 65 (47) |  |
| PDC, mean (SD) | 0.92 (0.13) | 0.92 (0.14) | 0.92 (0.11) | 0.93 (0.13) | 0.64 |
| Antidiabetic medications, N (%) | 84 (15) | 48 (17) | 19 (16) | 17 (12) |  |
| PDC, mean (SD) | 0.93 (0.09) | 0.93 (0.10) | 0.94 (0.09) | 0.96 (0.04) | 0.32 |
| Statin, N (%) | 173 (32) | 84 (29) | 43 (36) | 46 (33) |  |
| PDC, mean (SD) | **0.92 (0.10)** | **0.92 (0.10)** | **0.89 (0.13)** | **0.94 (0.06)** | **0.02** |

Abbreviations: PAP = positive airway pressure, PDC = proportion of days covered. Statistically significant findings are bolded.

Average (SD) hours of PAP use were 6.5 (1.0), 3.7 (1.2), and 0.5 (0.5) hours/night for the high, moderate, and low adherence groups, respectively. **Figure 1** and **e-Table 2** show the three groups’ average hours of PAP use for each six-month interval over the three years of follow-up. For the low and moderate adherence groups, average use was highest initially and decreased with time (*p*<0.001). On the other hand, the high adherence group started with high use and showed significantly increased use over time (p<0.001).

**Figure 1.**
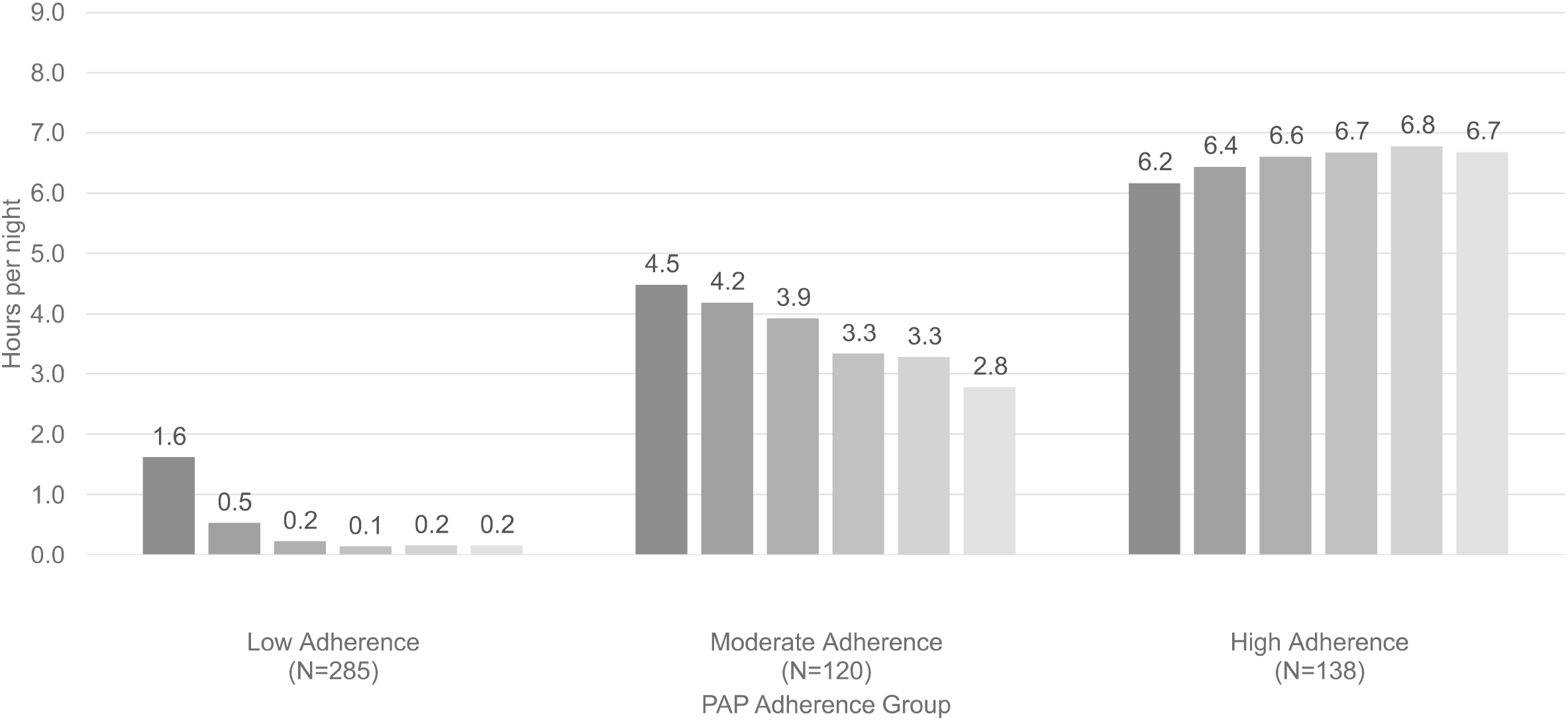
Mean Hours of PAP Use Over Time (Every 6 months) by PAP Adherence Abbreviations: PAP = positive airway pressure. Mo = Month. *p-*value for trend in each adherence group <0.001

Figure 2 and **e-Table 3** show average unadjusted healthcare costs over time. The six-month average cost prior to randomization (i.e., before OSA diagnosis and PAP initiation, represented by the left-most bar of each of the three clusters) was similar across the three adherence groups (*p=*0.91). The six-month average cost post to randomization in the high adherence group were significantly lower than those in the low adherence group (*p*=0.002) (**e-Table 3**), while differences between the moderate and low adherence groups (*p*=0.38) or between the high and moderate groups (*p*=0.08) were not statistically significant.

**Table 2** and **e-Table 4** show the adjusted average (SE) six-month costs by PAP adherence group for the overall sample as well as stratified by AHI. As with the unadjusted results, compared with the low adherence group, the high adherence group had significant adjusted six-month cost savings of $854 (95% CI ($158 to $1,551, *p*=0.01). Similarly, neither the adjusted difference in cost between the moderate and low adherence groups nor between the moderate and high adherence groups differed significantly.

**Table 2.** Adjusted Mean (SE) 6-Month Costs and Cost Savings Associated with Moderate or High PAP Adherence Stratified by Apnea Hypopnea Index (AHI)

|  | Adjusted Mean (SE)<br>Cost | Cost Savings (95% CI) from<br>[Moderate vs Low] or<br>[High vs Low] Adherence | Cost Savings (95% CI) from<br>[High vs Moderate] Adherence |
| --- | --- | --- | --- |
| **All** |  |  |  |
| Low | \$4,016 (\$315) | | |
| Moderate | \$3,658 (\$369) | \$359 (-\$459, \$1,176), *p*=0.40 | |
| High | \$3,162 (\$240) | **\$854 (\$158, \$1,551), *p*=0.01** | \$496 (-\$286, \$1,278), *p*=0.19 |
| **AHI: 5 to 14.9** |  |  |  |
| Low | \$4,170 (\$439) | | |
| Moderate | \$2,913 (\$350) | **\$1,257 (\$369, \$2,145), *p*=0.006** | |
| High | \$3,187 (\$354) | **\$983 (\$148, \$1,819), *p*=0.02** | -\$274 (-\$1,014, \$465), *p*=0.47 |
| **AHI: 15 or higher** |  |  |  |
| Low | \$3,665 (\$282) | | |
| Moderate | \$3,923 (\$429) | -\$258 (-\$1,155, \$639), *p*=0.56 | |
| High | \$3,087 (\$242) | \$578 (-\$123, \$1,279), *p*=0.11 | \$836 (-\$64, \$1,736), *p*=0.05 |

Abbreviations: PAP = positive airway pressure. Following covariates were included in the generalized linear models considering within-individual correlations and censoring weights: age, sex, race/ethnicity, body mass index, Medicaid, smoking status, hypertension, Charlson comorbidity, mild liver disease, prior 6-month cost, Apnea Hypopnea Index, pre-operative assessment, and time indicators (i.e. month 1-6, month 7-12, etc.). *p*-value for interaction (PAP adherence x AHI category) = 0.06. Statistically significant findings are bolded.

**Figure 2.**
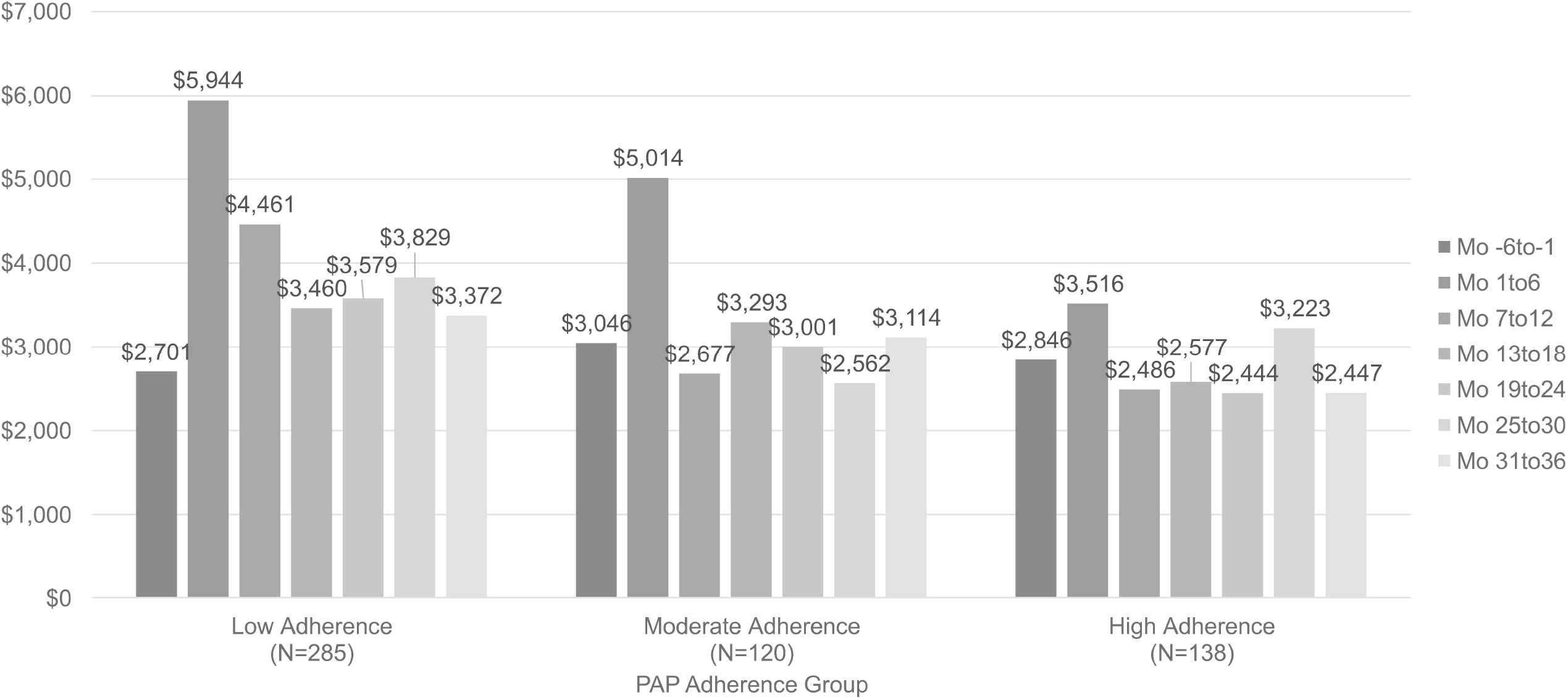
Mean Total Healthcare Costs Over Time (Every 6 months) by PAP Adherence Abbreviations: PAP = positive airway pressure. Mo = Month. *p-*value for trend in each adherence group <0.001

**Table 3** shows adjusted average six-month healthcare costs subdivided into 11 categories. The significantly lower overall cost in the high versus low adherence groups was due to significant savings in 5 of 11 cost categories: sleep-related office visit, non-sleep-related office visit, hospitalization, laboratory, and phone encounters while being slightly offset by a significant increase in sleep-related DME costs. The only three categories for which the high adherence group differed significantly from the moderate group were hospitalization, phone, and non-sleep-related DME. Finally, compared to the low adherence group, the moderate adherence group had significantly higher sleep-related DME costs.

**Table 3.**
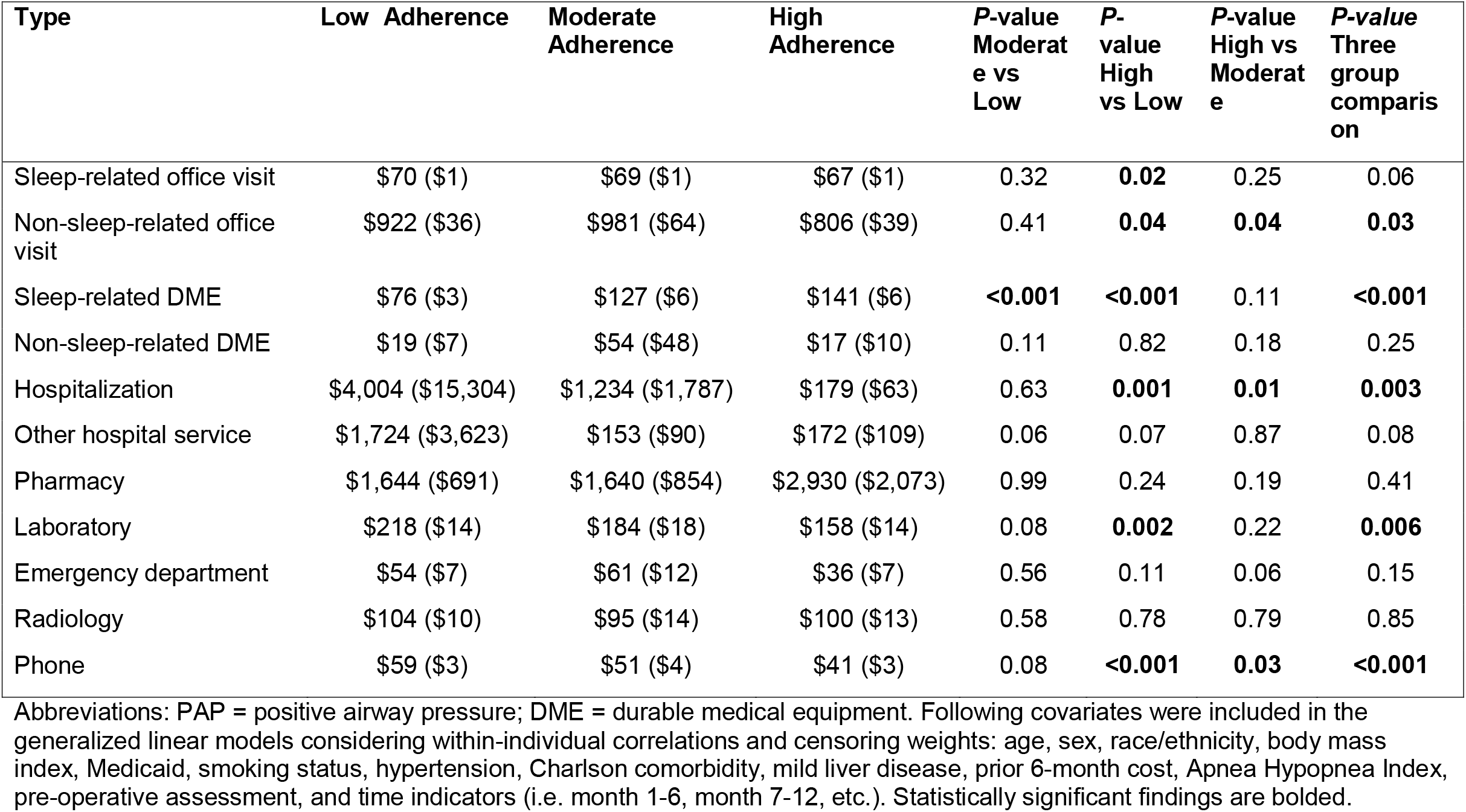
Adjusted Mean (SE) Six-Month Healthcare Costs by Type of Healthcare Use and PAP Adherence.

| Type | Low Adherence | Moderate Adherence | High Adherence | *P*-value Moderate vs Low | *P*-value High vs Low | *P*-value High vs Moderate | *P*-value Three group comparison |
| --- | --- | --- | --- | --- | --- | --- | --- |
| Sleep-related office visit | \$70 (\$1) | \$69 (\$1) | \$67 (\$1) | 0.32 | **0.02** | 0.25 | 0.06 |
| Non-sleep-related office visit | \$922 (\$36) | \$981 (\$64) | \$806 (\$39) | 0.41 | **0.04** | **0.04** | **0.03** |
| Sleep-related DME | \$76 (\$3) | \$127 (\$6) | \$141 (\$6) | **<0.001** | **<0.001** | 0.11 | **<0.001** |
| Non-sleep-related DME | \$19 (\$7) | \$54 (\$48) | \$17 (\$10) | 0.11 | 0.82 | 0.18 | 0.25 |
| Hospitalization | \$4,004 (\$15,304) | \$1,234 (\$1,787) | \$179 (\$63) | 0.63 | **0.001** | **0.01** | **0.003** |
| Other hospital service | \$1,724 (\$3,623) | \$153 (\$90) | \$172 (\$109) | 0.06 | 0.07 | 0.87 | 0.08 |
| Pharmacy | \$1,644 (\$691) | \$1,640 (\$854) | \$2,930 (\$2,073) | 0.99 | 0.24 | 0.19 | 0.41 |
| Laboratory | \$218 (\$14) | \$184 (\$18) | \$158 (\$14) | 0.08 | **0.002** | 0.22 | **0.006** |
| Emergency department | \$54 (\$7) | \$61 (\$12) | \$36 (\$7) | 0.56 | 0.11 | 0.06 | 0.15 |
| Radiology | \$104 (\$10) | \$95 (\$14) | \$100 (\$13) | 0.58 | 0.78 | 0.79 | 0.85 |
| Phone | \$59 (\$3) | \$51 (\$4) | \$41 (\$3) | 0.08 | **<0.001** | **0.03** | **<0.001** |

Abbreviations: PAP = positive airway pressure; DME = durable medical equipment. Following covariates were included in the generalized linear models considering within-individual correlations and censoring weights: age, sex, race/ethnicity, body mass index, Medicaid, smoking status, hypertension, Charlson comorbidity, mild liver disease, prior 6-month cost, Apnea Hypopnea Index, pre-operative assessment, and time indicators (i.e. month 1-6, month 7-12, etc.). Statistically significant findings are bolded.

An interaction test examining whether the cost savings associated with specific adherence groups differed based on AHI severity was non-significant (*p*=0.06). However, given the limited power of this test, we also performed an *a priori* specified stratified analysis. Results showed that among participants with mild OSA (AHI 5-14.9), the high and moderate adherence groups both had significant savings compared to the low group ($938, 95% CI $148 to $1,819, *p*=0.02 for high vs low and $1,257, 95% CI $369 to $2,145, *p*=0.006 for medium vs low). There were, however, no significant differences between the high and moderate groups. Unlike the findings for those with mild OSA, among participants with moderate to severe OSA (AHI ≥ 15), there were no statistically significant differences between any of the three adherence groups. Finally, in sensitivity analysis, findings were consistent after the exclusion of participants who were referred to sleep clinic prior to planned surgery (see **e-Table 4**).

## Discussion

The current study evaluated healthcare costs associated with PAP adherence among participants in the Tele-OSA clinical trial. Using daily PAP use data over a three-year follow-up period, this study stratified participants into high, moderate, and low long-term adherers to PAP therapy. The integrated healthcare system with electronic health records captured all aspects of care; therefore, we were able to evaluate the relationship between PAP adherence and healthcare costs across a broad range of services. Our results demonstrated lower healthcare costs associated with high adherence compared with low adherence supporting the potential economic benefits of PAP therapy.

Although OSA is understood to impact healthcare costs through its association with cardiovascular disease, mental health disorders, peri-operative risk, and rate of hospitalizations [8, 19, 59]., there has been lack of economic evaluation studying the potential impact of PAP use. Furthermore, the few previous studies have not consistently reported a beneficial association between PAP therapy and cost. An earlier study from the Kaiser Permanente healthcare delivery system failed to demonstrate a significant impact of PAP dispensation on acute care and pharmacy utilization, although PAP adherence was not available to be included in the analysis [46]. A recent study by Kirsch *et al*. reported that use of PAP at least 4 hours/night over an 18-month period in participants with moderate-severe OSA was associated with lower inpatient visits and costs compared with non-adherers [44]. Our study adds to the body of evidence that long-term adherence to PAP therapy can beneficially impact healthcare costs.

Several interesting patterns emerged from this study that warrant further investigation. First, mimicking the findings in the Kirsch et al study, savings appeared to be driven by reduced cost related to hospitalizations. Further understanding the specific services for which there are savings from PAP therapy carries potential implications in developing administrative and clinical care strategies. Second, PAP use among participants with mild OSA was observed to result in greater cost savings than among those with moderate-severe OSA. While this apparent reverse dose-response pattern should be replicated within larger samples, the results are supportive of the hypothesis that treating participants with mild OSA may be clinically important. Future confirmation of this finding could influence approaches to clinical care and development of research cohorts that currently tend to focus on moderate-severe OSA. Third, PAP adherence stratification was based not only on absolute usage but also on the consistency of maintaining adherence over time. Cost reductions realized among high adherers but not clearly evident among moderate adherers are suggestive of the hypothesis that both sufficient and consistent use are important to improve outcomes. Our data also highlight the challenge of meeting both adherence targets—only 25% of the analysis cohort used PAP consistently ≥ 4 hours/night over three years. Further understanding the influence of PAP adherence patterns on healthcare outcomes is important in strategizing effective clinical care follow-up protocols.

This study has several potential limitations. First, although we adjusted for participant sociodemographic and clinical characteristics as covariates, there may be unmeasured confounders. In particular, healthy adherer bias is a typical concern in the adherence literature [60-63]; participants adherent to PAP therapy may also be more likely to adhere to other medical care (e.g., medication use or receiving preventative therapy) or healthy lifestyle behaviors (e.g., diet and exercise). However, our assessment of medication adherence at baseline as well as six-month healthcare costs prior to randomization were similar across PAP adherence groups. Second, our study population was comprised of clinical trial participants which could limit generalizability to real-world populations. However, the Tele-OSA study was conducted in a pragmatic setting that enrolled participants at risk for OSA typical of a standard sleep medicine practice, which helps mitigate the potential limits in generalizability. Third, there may be misclassification bias [64]. We used diagnosis codes from electronic health records to define comorbidities; therefore, comorbidities of participants who did not interact with the healthcare system or participants without continuous eligibility may have not been fully captured. However, 86% of the study population had continuous eligibility for 12 months prior to the index date, and all participants had at least one interaction with the healthcare system. Fourth, we evaluated healthcare costs by assigning nationally representative costs instead of paid claims. However, this approach is recognized as a generalizable method to other healthcare settings [58, 65].

## Interpretation

Better PAP adherence (consistent PAP use of ≥ 4 hours/night) was associated with significantly lower healthcare costs over three years in participants with OSA. Findings support the importance of care strategies to enhance long-term PAP adherence for OSA therapy.

## Data Availability

Anonymized data that support the findings of this study may be made available from the investigative team with the following conditions: 1) agreement to collaborate with the study team on all publications, 2) provision of external funding for administrative and investigator time necessary for this collaboration, 3) demonstration that the external investigative team is qualified and has documented evidence of training for human subjects protections, and 4) agreement to abide by the terms outlined in data use agreements between institutions.

## Data Availability

All data related to this work is available upon request

## Guarantor

Hwang takes responsibility for the content of the manuscript, including the data and analysis

## Author Contributions

An contributed to the study design, data analysis, interpretation, and initial draft of the manuscript. Glick, Sawyer contributed substantially to the study design, data analysis and interpretation, and the writing of the manuscript. Arguelles, Chen, Shi contributed data acquisition, programming, data analysis, and the critical revision of the manuscript. Bae, Keenan, Kuna, Maislin, Mazzotti, Pack, Watach contributed to the study design, interpretation, and the critical revision of the manuscript. Hwang had full access to all of the data in the study and takes responsibility for the integrity of the data and the accuracy of the data analysis.

## Prior Abstract Publication/Presentation

Preliminary findings were presented as a poster at Virtual Sleep, 2021 (June 10-13, 2021). An J, Glick H, Shi J, Chen A, Arguelles J, Keenan B, Maislin G, Mazzotti D, Sawyer A, Pack A, Hwang D. Association between Positive Airway Pressure Adherence and Healthcare Costs, Sleep, Volume 44, Issue Supplement_2, May 2021, Page A177, https://doi.org/10.1093/sleep/zsab072.447

## Abbreviation List

AHI: Apnea Hypopnea Index
ANOVA: analysis of variance
BMI: body mass index
CPT: Current Procedural Terminology
DME: durable medical equipment
GLM: generalized linear models
HCPCS: Healthcare Common Procedure Coding System
NADAC: National Average Drug Acquisition Cost
OSA: Obstructive sleep apnea
PAP: positive airway pressure

